# Taking on colonial institutions: making room for an Indigenous Research Paradigm in prospective biomedical research in tertiary hospitals

**DOI:** 10.1101/2024.08.06.24311521

**Authors:** Jessica O’Brien, Sarah J Gutman, Vicki Wade, Toni Walker, Andrew J Taylor, Karen Adams

**Author notes:** **Address for correspondence:** Professor Karen Adams Gukwonderuk Indigenous Health Unit Faculty of Medicine, Nursing and Health Sciences Monash University Wellington Road, Clayton, Victoria, Australia 3800.

## Abstract

**Background:** A paradigmatic clash exists between biomedical and Indigenous research frameworks. Problematically, when ill-fit biomedical research frameworks are applied Indigenous peoples can experience exclusion from biomedical studies and consequent potential health benefits. To overcome these issues, community based participatory research methodologies are often recommended. However, these can prove difficult to apply in tertiary healthcare research where prospective Indigenous peoples and families participating in research will come from unforeseen and numerous Indigenous communities. Adding further complexity, there appears a dearth of information for achieving incorporation of decolonising and Indigenous research frameworks into this type of prospective research.

**Methods:** We sought to reflect on and describe inclusion of an Indigenous Research Paradigm into the establishment of a prospective multi-site tertiary healthcare study on improving diagnosis and treatment of acute rheumatic fever. To generate reflection, the First Nations’ Yarning method was employed allowing qualitative findings to be generated via Indigenous epistemology and ontology.

**Findings:** Four main areas were identified as requiring significant change to align with an Indigenous Research Paradigm: stakeholder engagement, project design, consent processes, and multi-site approach. Multi-layered Indigenous leadership was recognised as a crucial component of the transformation and of the project’s success more broadly.

**Interpretation:** With extensive local First Nations involvement and a First Nations-led research team, a multi-site institution-based biomedical research project can be successfully adapted to be more in keeping with an Indigenous Research Paradigm.

## Background

Worldwide concerns exist that biomedical research is being performed on Indigenous peoples (meaning Indigenous peoples globally) rather than with, for or by us.^1^ Consequently Indigenous peoples stipulate inclusion of Indigenous leadership and knowledges in biomedical research as crucial.^1–3^ Community-based participatory research (CBPR) methods are often recommended to facilitate inclusion.^4^ These methods are particularly useful as Indigenous research is commonly locally grounded involving “politics, circumstances, and economies of a particular moment, a particular time and place, a particular set of problems, struggles, and desires” ^5, p.14^. The CBPR methods involve researchers and community members as equal partners throughout the research, combining research with community empowerment to generate relevant, meaningful, and responsive benefits for that community.^6^ However, CBPR methods can prove difficult to apply in tertiary healthcare research where prospective Indigenous peoples and families participating in research will come from numerous and unforeseen Indigenous communities.

Due to ill-fit research methodologies, Indigenous peoples can experience exclusion from prospective tertiary healthcare research and consequent potential health benefits. For instance, exclusion may be driven by researchers: imposing culturally unsafe research methods upon Indigenous peoples or; deeming culturally safe prospective research too difficult to achieve. As such, an equity and human rights imperative exists to improve and innovate prospective tertiary healthcare research methodology. Here we describe the responsive pivots taken to engage an Indigenous Research Paradigm in the establishment of a multi-site hospital-based prospective study examining the role of cardiac magnetic resonance imaging (MRI) in diagnosing acute rheumatic fever (ARF), a condition that inequitably affects First Nations peoples (meaning Aboriginal and Torres Strait Islander people) in Australia.^7^

### A biomedical and Indigenous research paradigmatic clash

This research is underpinned by what Nakata^8^ theorises as a cultural interface. This interface recognises that Indigenous and colonial knowledge systems exist and that these are so unalike that there is little to no commonality.^8^ In accordance with this understanding, this research is deeply rooted in a paradigmatic clash between colonially informed biomedical research and an Indigenous Research Paradigm. An Indigenous Research Paradigm: values relationality and relational accountability;^9^ is driven by Indigenous research interests; is holistic;^1^ involves Indigenous perspective to choose methods and decide what is knowledge is^10^ and; creates emancipation and benefit for Indigenous peoples.^11^ In contrast, Biomedical Research Paradigms can be understood as: embedded in objectivism: seeing knowledge as awaiting discovery; valuing positivism and empiricism; forming a tight grid around what it observes^12^ and; favouriting individualism.^13^ As such, Indigenous and Biomedical Research Paradigms are very distinct and dissimilar. However, there is an additional socio-cultural-historic layer to this paradigmatic clash as the Biomedical Research Paradigm has been applied as a colonial tool of oppression.^14^

Historically biomedical researchers created discredited and harmful racialized theories which supported destruction of Indigenous peoples lives and livelihoods via colonisation.^15^ It is tempting to consider these ideas as something of an antiquated past, however, colonisation is a shapeshifter^16^ and the past is inextricably linked to the present and making of the future. For instance, researchers can study health inequities imposed upon Aboriginal peoples with little self-awareness of their own profession’s complicity in causing the disparities in the first place. The forgetting of the past likely informs the troubling tendency of biomedical research to infer that Aboriginal peoples are inherently deficient. Some have described this as a deficit discourse^17^ or as the 5Ds of Indigenous representation in research - difference, disparity, disadvantage, dysfunction, and deprivation.^18^ Similar to yesteryear, Indigenous peoples are subtly blamed for health inequities and consigned to being weaker or lesser than non-Indigenous people, particularly when, the socio-cultural-historic factors causing the inequities are absented. It is within this complex and contested interface between divergent paradigms that consideration was given to the establishment of a prospective biomedical research study.

## Methods

### Research aim

We sought to reflect on and describe inclusion of an Indigenous Research Paradigm into the establishment of a prospective multi-site tertiary healthcare study on improving diagnosis and treatment of ARF.

### Study context

ARF is a generalised inflammatory illness that occurs following Group A Streptococcal infection.^19^ As in many other colonised high-income countries, ARF has been essentially eradicated in Australia’s non-Indigenous population, but endemic rates continue in children and young people in many First Nations communities,^20,21^ predominantly in regional, remote and very remote areas. ^7^ This imposed inequity is the result of ongoing colonisation upon First Nations peoples which has created a situation where ARF thrives on poverty, social disadvantage, and poor healthcare access.^22^ (See Figure 1). If ARF is not recognised and further episodes prevented with monthly intramuscular penicillin injections, rheumatic heart disease (RHD) and its complications can occur.

**Figure 1.**
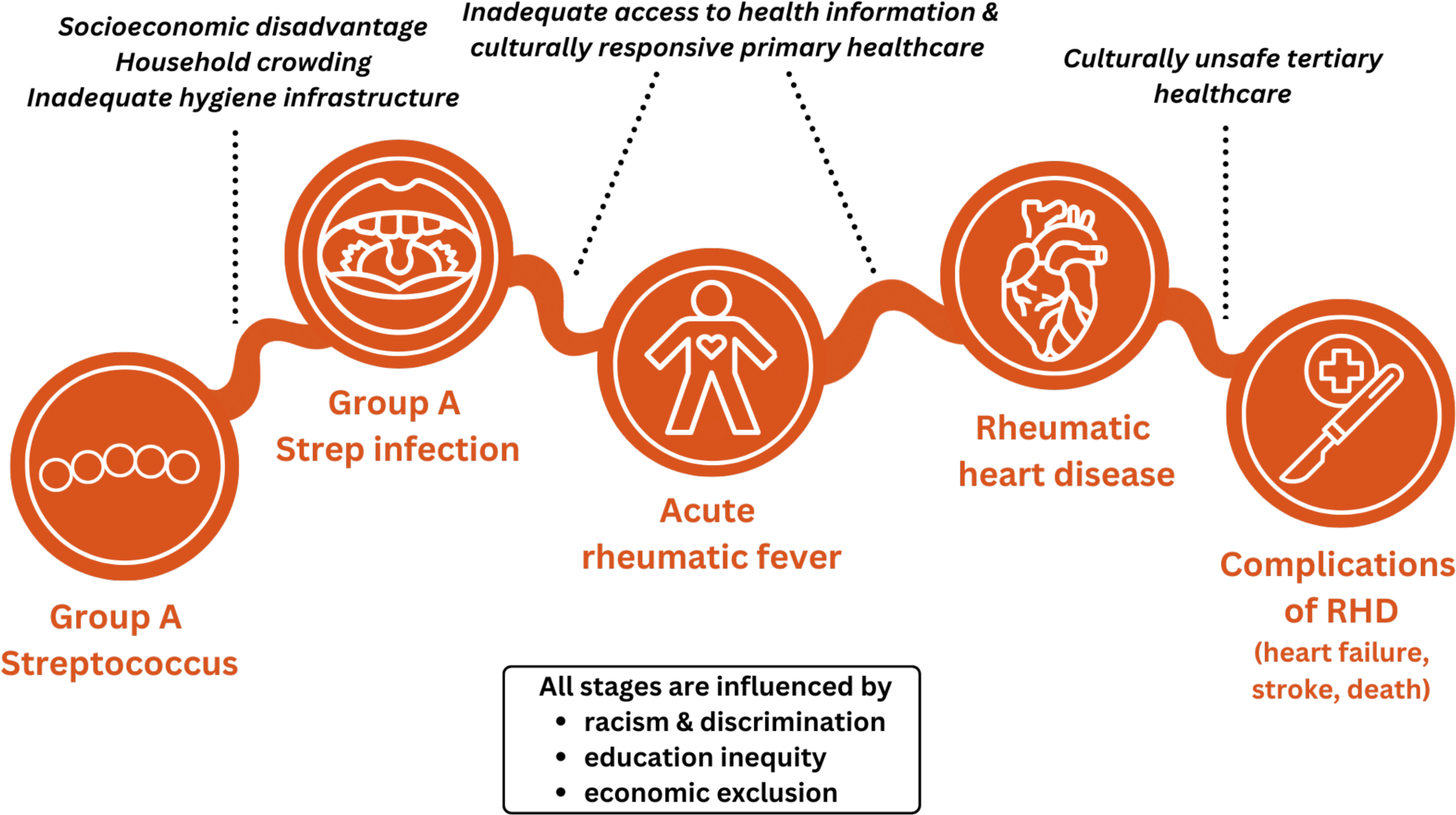
**Social determinants of health and progression to RHD** The social determinants of health influence progression of Group A Streptococcus to RHD. Culturally responsive primary and tertiary healthcare is crucial in halting this progression. RHD = rheumatic heart disease

In Australia, when ARF is diagnosed or suspected, patients are transferred to tertiary hospitals for further investigations, assessment by a multi-disciplinary specialist team, as well as education about ARF and RHD. Diagnosis is based on a combination of clinical manifestations and several investigations; there is no single diagnostic test.^7^ As a result, “possible” and “probable” ARF categories have been developed to ensure mild and atypical presentations are not missed within high-risk populations.^7^ This categorisation guides the duration of secondary antibiotic prophylaxis, the only proven way of controlling RHD at the population level.^23^ However, the efficacy of secondary prophylaxis is balanced against the significant burden it places on patients, families, communities, and health services.

The ARF and RHD inequities informed the establishment of a project entitled ‘Quantifying myocardial inflammation in ARF and RHD’ or ‘I Heart MRI’ (see Appendix 1 for full study protocol and figure 2 for protocol summary). It involves participants with ARF, non-ARF inflammatory conditions and those with no known cardiac or inflammatory issues (healthy controls) undergoing a truncated non-contrast cardiac MRI scan during admission to identify and quantify myocardial inflammation. Data from three participating sites, Alice Springs, Cairns, and Darwin hospitals will be combined to determine if a diagnostic score for ARF can be developed, thereby clarifying who will benefit most from secondary prophylaxis.

**Figure 2.**
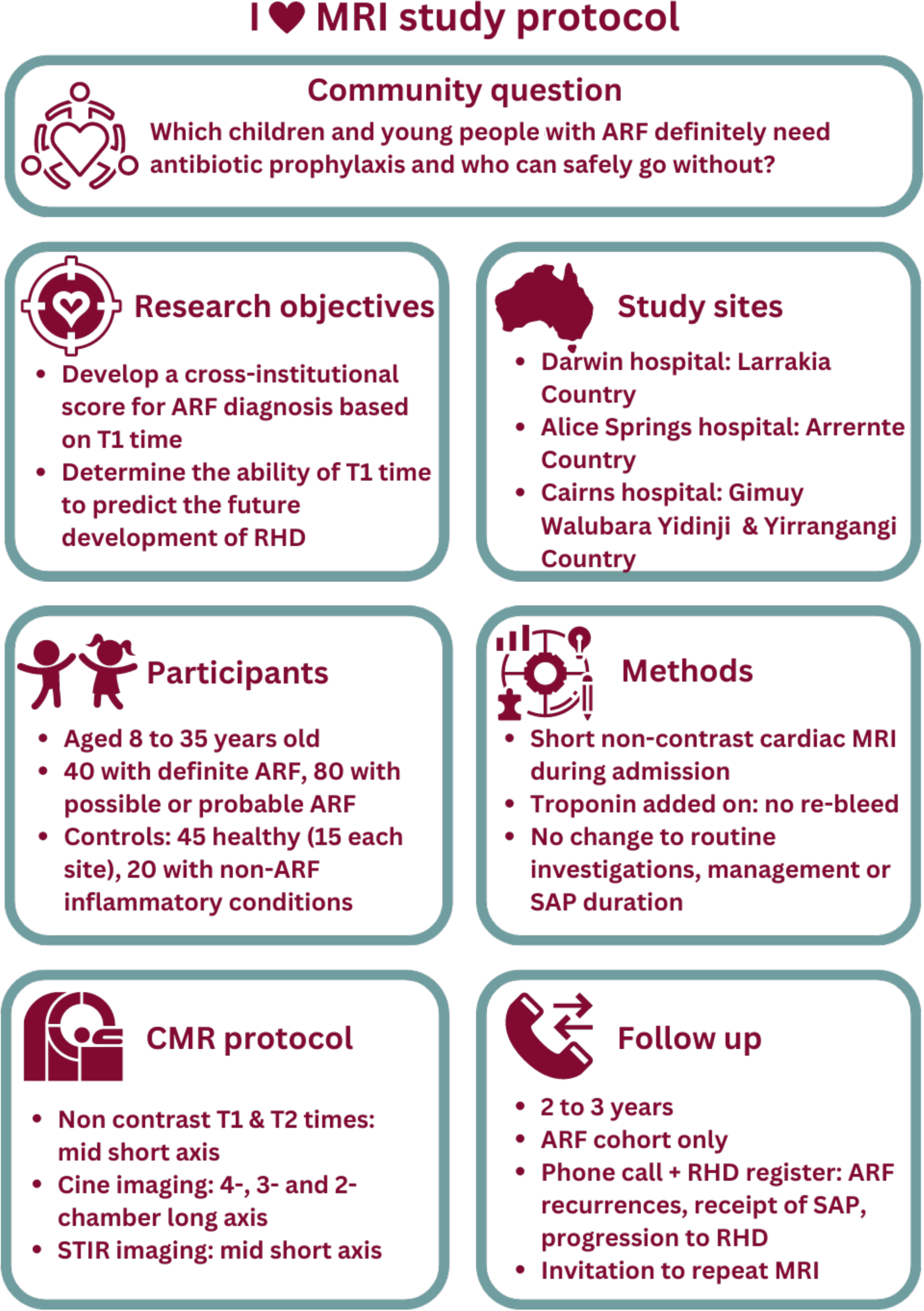
**I Heart MRI study protocol summary** Caption: Summarised study protocol for I Heart MRI MRI = magnetic resonance imaging; ARF = acute rheumatic fever; RHD = rheumatic heart disease; SAP = secondary antibiotic prophylaxis

Participants in the ARF group will be followed-up at two to three years to determine if they have progressed to RHD. The establishment of the study took place over a three-year period with recruitment commencing April 2023.

### Researcher positionality

An important aspect of Indigenous and decolonising research is to declare standpoint. This involves researchers stating who they are and considering how this positions them in society and in relation to participants and the research itself.^10^ Positionality provides context to the research that is important for participants and users of the research to know.^24^ Related to this is reflexivity, the process of researchers actively and continuously assessing their potential biases and how these impact upon assumptions, intentions, decisions, and reactions.^24^ Both positionality and reflexivity positively influence research design and interpretation of results, and strengthen participant-researcher relationships.

JO is an Aboriginal woman from northwest New South Wales, a cardiologist trained in a traditional biomedical system with limited experience in conducting research with First Nations peoples prior to leading this project as part of her PhD. Her insider status to participants as an Aboriginal person with relatives with ARF and RHD is balanced against being an outsider as a privileged medical professional living in a highly urbanised area where rates of ARF are extremely low. The supervising author, KA, is Wiradjuri and has worked in Aboriginal health in Southeast Australia as a nurse, health service manager, educator and researcher for more that 25 years. Other authors did not participate in the Yarning but contributed to the design and establishment of ‘I Heart MRI’ as well as to manuscript production. VW is a senior Noongar woman and cardiac nurse with decades of clinical and research experience in ARF and RHD. TW is Gurang and has worked as a senior Aboriginal Health Practitioner for two decades across Queensland and the Northern Territory. SJG and AJT are non-Indigenous academic cardiologists with expertise in cardiac MRI.

### Collaborative Yarning method

When generating Indigenous knowledges, the process for doing this is relevant and crucial.^1^ For instance, constructing knowledge with processes underpinned by Indigenous epistemologies and ontologies will cyclically allow for further expression of Indigenous epistemologies and ontologies to be generated. Therefore, this research engaged Yarning method, a First Nations culturally specified communication process.^25^ Yarning method involves people coming together with a purpose and is relationally based.^26^ Therefore, the existing tacit and explicit experiences and memories that people have in common to inform relationality is important, as, this will impact data thickness and quality.^25^ The Yarning method can involve different types of Yarning such as: Social Yarning to build relationship; Research Yarning to gather information and stories; Collaborative Yarning to share research information, discuss ideas and develop new concepts and; Therapeutic Yarning whereby research participants can tell their story and make greater sense of it.^26^

In this study, the Yarning purpose was to engage in a series of Yarns to gather stories about development of a research project engagement methodology; reflect on these stories to make sense of them and develop ideas and concepts. Six yarns 1-1.5 hours long took place between February and August 2023 involving Research, Collaborative and Therapeutic Yarns (see Table 1). The authors took notes during Yarns and identified responsive pivots that were important to establishing the study. As these are personal reflections of the authors ethics approval was not sought for the Yarning. However, the broader ARF study has ethics approval provided by the Far North Queensland Human Research Ethics Committee (HREC) with project ID 67038-1493 and the HREC of the Northern Territory Department of Health and Menzies School of Health Research with project ID 2020-3887.

**Table 1.**
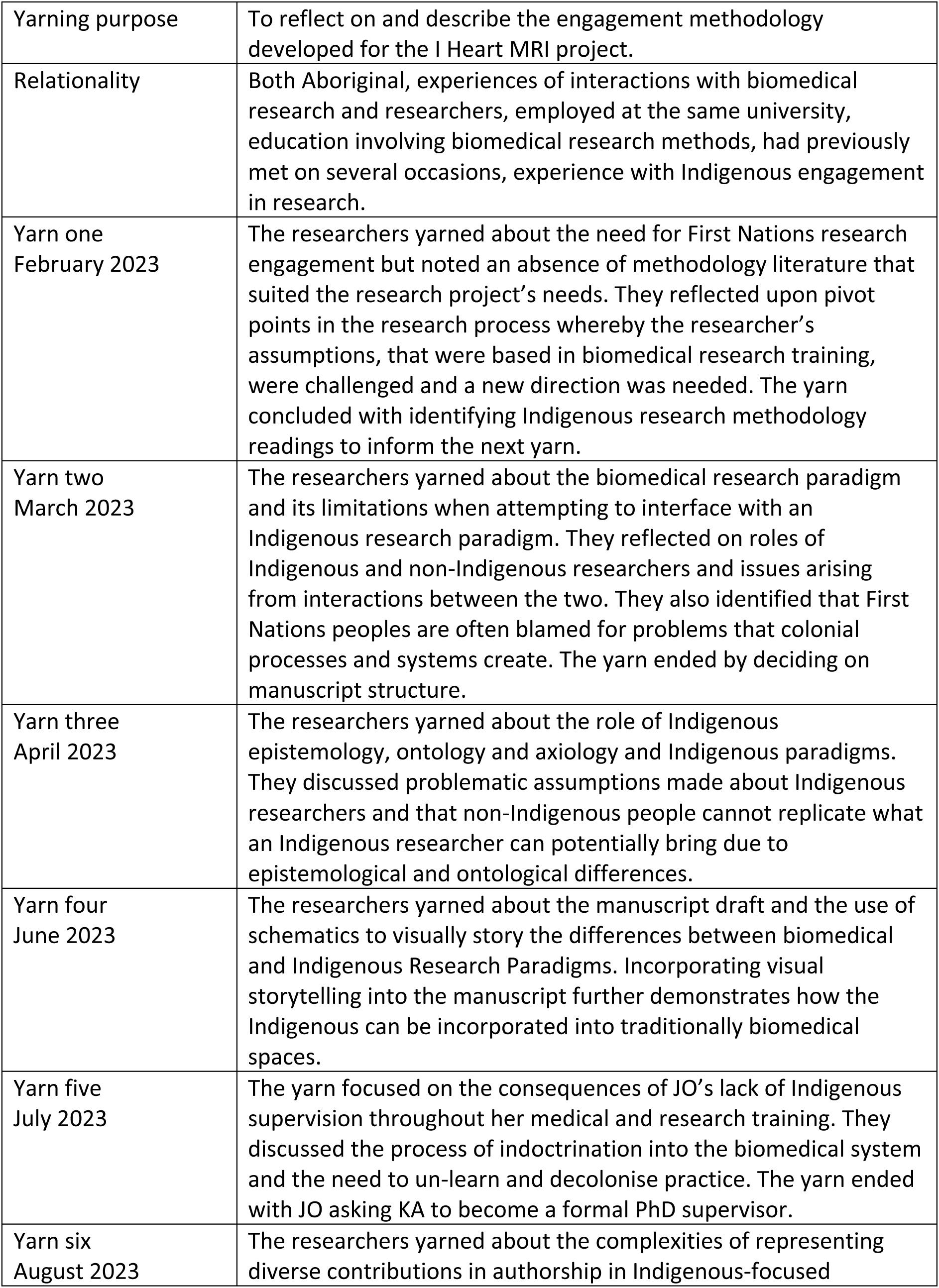
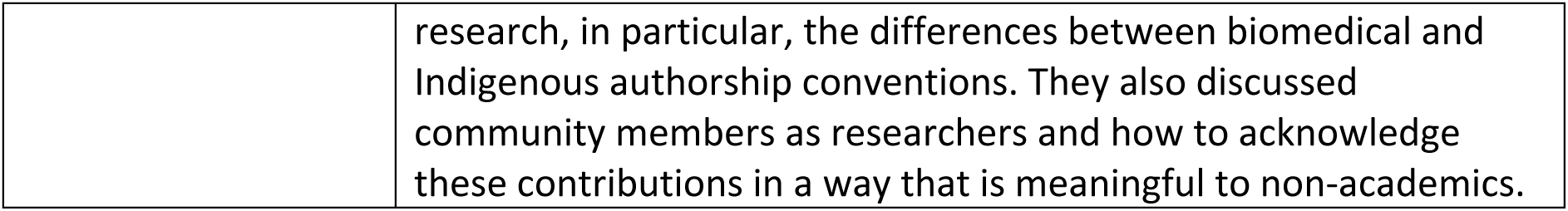
Summary of collaborative Yarning process between Professor Karen Adams and Dr Jessica O’Brien.

|  |  |
| --- | --- |
| Yarning purpose | To reflect on and describe the engagement methodology developed for the I Heart MRI project. |
| Relationality | Both Aboriginal, experiences of interactions with biomedical research and researchers, employed at the same university, education involving biomedical research methods, had previously met on several occasions, experience with Indigenous engagement in research. |
| Yarn one<br>February 2023 | The researchers yarning about the need for First Nations research engagement but noted an absence of methodology literature that suited the research project's needs. They reflected upon pivot points in the research process whereby the researcher's assumptions, that were based in biomedical research training, were challenged and a new direction was needed. The yarn concluded with identifying Indigenous research methodology readings to inform the next yarn. |
| Yarn two<br>March 2023 | The researchers yarning about the biomedical research paradigm and its limitations when attempting to interface with an Indigenous research paradigm. They reflected on roles of Indigenous and non-Indigenous researchers and issues arising from interactions between the two. They also identified that First Nations peoples are often blamed for problems that colonial processes and systems create. The yarn ended by deciding on manuscript structure. |
| Yarn three<br>April 2023 | The researchers yarning about the role of Indigenous epistemology, ontology and axiology and Indigenous paradigms. They discussed problematic assumptions made about Indigenous researchers and that non-Indigenous people cannot replicate what an Indigenous researcher can potentially bring due to epistemological and ontological differences. |
| Yarn four<br>June 2023 | The researchers yarning about the manuscript draft and the use of schematics to visually story the differences between biomedical and Indigenous Research Paradigms. Incorporating visual storytelling into the manuscript further demonstrates how the Indigenous can be incorporated into traditionally biomedical spaces. |
| Yarn five<br>July 2023 | The yarn focused on the consequences of JO's lack of Indigenous supervision throughout her medical and research training. They discussed the process of indoctrination into the biomedical system and the need to un-learn and decolonise practice. The yarn ended with JO asking KA to become a formal PhD supervisor. |
| Yarn six<br>August 2023 | The researchers yarning about the complexities of representing diverse contributions in authorship in Indigenous-focused |
|  | research, in particular, the differences between biomedical and Indigenous authorship conventions. They also discussed community members as researchers and how to acknowledge these contributions in a way that is meaningful to non-academics. |

## Results

Collaborative Yarning identified four key responsive pivots requiring research design transformation to alter a Biomedical research approach to align better with an Indigenous Research Paradigm (see Figure 3).

**Figure 3.**
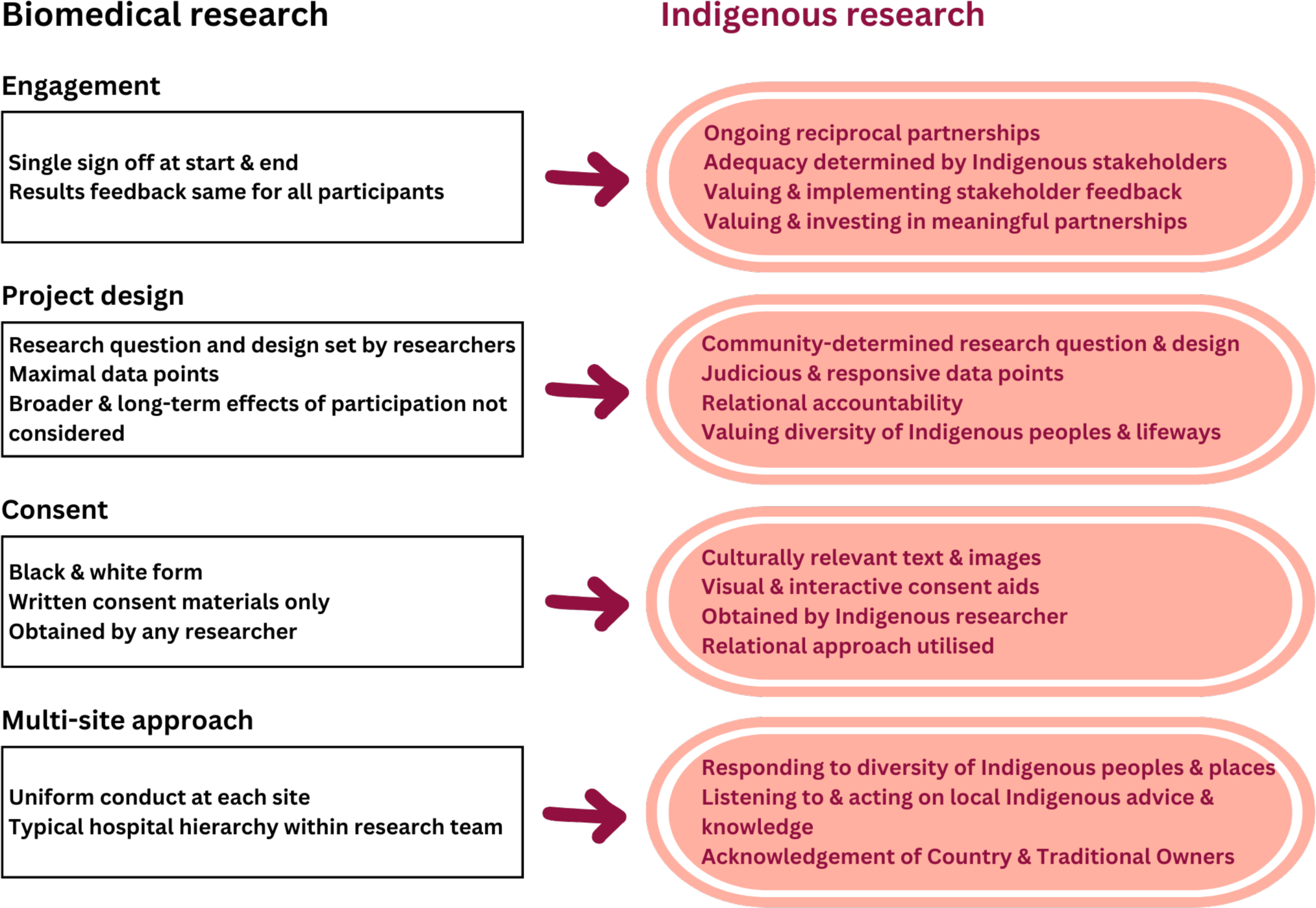
**Biomedical to Indigenous Research Paradigm pivot points** Four major aspects of the project underwent significant change to be more in keeping with an Indigenous Research Paradigm

### Pivot One – Stakeholder Engagement

#### Perceived Research Paradigm Clash

Biomedical research designs typically involve development of a research question followed by engagement with stakeholders. Ethics and governance committees commonly decide if sufficient engagement has occurred, and it often happens only at the start and end of a project. Methods of providing feedback of results to First Nations peoples can be unchanged from those used for non-Indigenous participants and stakeholders, or feedback processes can be omitted entirely. In contrast, First Nations research ethics guidelines recognise that research questions require development with First Nations peoples and engagement requires ongoing reciprocal relationships with multiple stakeholders ^27^ (see Figure 4). Furthermore, First Nations peoples, in collaboration with researchers, determine: whether engagement is sufficient; the conditions under which they will allow the research to proceed; what is expected going forward and; what is required in return for the knowledge, time and effort given.^27^

**Figure 4.**
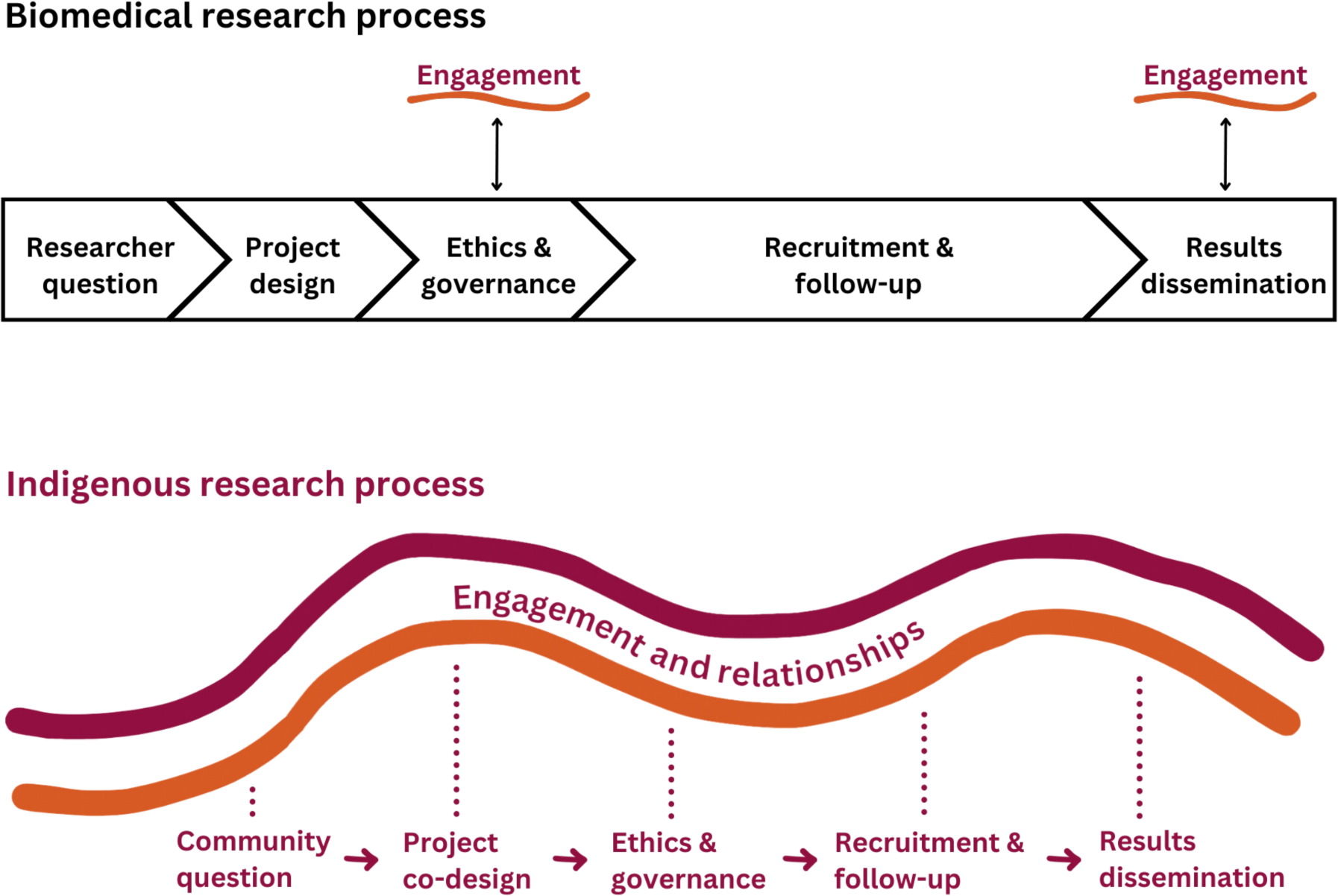
**Biomedical and Indigenous Research engagement** In a biomedical paradigm, the project is central with engagement occurring at time points that meet researchers needs. In an Indigenous paradigm, engagement is central, meaningful and enduring, with the project progressing as determined by community.

#### Responsive Pivots

Initially I Heart MRI utilised a typical top-down biomedical approach with the research question, design and funding established before First Nations stakeholder opinions were sought. Knowing where to start with community engagement at each site was difficult with little to no relationality established. The top-down approach made engagement more difficult as the methods the researchers suggested proved an ill-fit to each site. While this error was rectified, it delayed project set-up and unnecessarily burdened multiple First Nations stakeholders. A better fit was pivoting to Yarning with First Nations employees at each hospital to learn about: the different mobs (First Nations peoples) admitted; how the project could best be conducted; and who needed to be engaged in the research. Employing relational accountability, author one contacted every person, committee and organisation suggested by the First Nations employees. The suggested contacts proved crucial and provided introduction to additional key First Nations stakeholders such as Community Controlled Organisations (CCOs).

Essential to engaging CCOs was showing respect and reciprocity by following CCO advice regarding appropriate engagement methods. Particularly as the project establishment occurred during the Covid-19 pandemic and CCOs had limited time to support a predominantly hospital-run study. However, most CCOs agreed it was imperative they knew about the study. The CCO-requested engagement methods included information about the research alongside education sessions about ARF and other cardiac conditions. This ensured education went beyond a niche research project to more relevant topics for holistic primary healthcare.

A further critical stakeholder group engaged were families impacted by ARF. By Yarning with this all-important group, author one gained better understanding of the health consumer realities of diagnosis and admission experiences. The families provided crucial advice for consent processes and how to strengthen cultural safety and age appropriateness (see consent responsive pivot) for engaging with MRI. Additionally, a First Nations Advisory Group provides oversight for the project. The group consists of seven members, all First Nations peoples and of varied backgrounds in health and research. Furthermore, the First Nations representatives of national stakeholder organisations like RHD Australia and the Heart Foundation were engaged early to ensure prompt translation of study results to clinical practice, and therefore, tangible benefit for First Nations peoples. Liaising with these organisations via First Nations cultural leads rather than their biomedical counterparts was a conscious decision to mark this project as First Nations led and centred from the outset. First Nations leads also advocated for engagement of other institutions performing research at the same sites to coordinate approaches to recruitment so that potential participants were not burdened by repeated consent attempts.

An additional pivot involved changing the intended method of results feedback to stakeholders. A single page of lay summary results with the same page provided to each of the three sites was changed to a summary tailored to each site. These will further be informed by local First Nations perspectives with graphs and illustrations used to story results visually where possible. Short videos will also be created and made available to participants, CCOs, RHD program staff, First Nations hospital employees and any other groups routinely involved in the care of those with ARF and RHD.

### Pivot two – project design

#### Perceived Research Paradigm Clash

A biomedical approach typically values establishing a research design that predominantly revolves around the researcher’s needs to collect robust data. In contrast, an Indigenous Research Paradigm additionally considers local contexts and is relationally responsive to local needs.

#### Responsive Pivots

Initially the I Heart MRI study involved a troponin blood test, longer cardiac MRI with more sequences and many questions in the participant survey to maximise data collection. Two-year follow-up was to include a questionnaire, echocardiogram and repeat MRI for all participants in the ARF group. Upon engagement feedback the project design pivoted to ensure the research question was still answered in a scientifically robust manner, but in a way that was responsive to local contexts and needs. Of preeminent importance was providing a least invasive and non-burdening healthcare experience. This was particularly crucial to establish and maintain positive ongoing relationships to healthcare.

As such, only non-contrast MRI sequences are conducted removing the need for cannulation. If not already assessed, troponin assays are run on previously collected blood rather than performing repeat venesection. A repeat MRI for all at the arbitrary two-year mark was amended as it was felt that the information to be obtained did not justify burdening participants to attend often geographically distant institutions. Data from a smaller number of participants who live closer to the hospital, tolerated the baseline MRI well or who have particular interest in the project can be used to determine this secondary endpoint instead. However, all participants will be offered the choice and those declining will not be given a deficit label of “lost to follow-up” or “drop out”.

Finally, a follow-up research echocardiogram was abandoned in favour of using locally performed clinical echocardiograms which will adequately determine whether participants have progressed to RHD. Engagement feedback identified a further required pivot. This related to some patients being likely to change addresses or phone numbers and experience numerous other barriers to attending appointments. Thus, participants will be contacted for the follow-up questionnaire, however, where participants cannot be contacted, the Northern Territory and Queensland RHD registers will be used to access information about antibiotic prophylaxis and echocardiogram results. Permission for the team to access register information is part of the research consent process.

### Pivot point three – consent

#### Perceived research paradigm clash

Often biomedical consent processes are minimally relational and focus on achieving individual consent. In contrast within an Indigenous Research Paradigm relationality in consent is highly valued.

#### Responsive pivots

Initially this research conceptualised consent as involving a template consent form to be signed by the participant or guardian following discussion about the project with a research team member. We pivoted to engaging Yarning and relationality with a starting focus on cultural and language background of patients and families. The researcher also shares who they are and their role in the research. Where possible, consent is undertaken by First Nations team members. Potential participants are also offered involvement of a First Nations liaison officer, health worker or interpreter in consent Yarns. This aims to add comfort by increasing non-biased support and the number of First Peoples present. Family or community members can also be invited to consent processes via telephone or video conference. These processes often lead to multiple discussions prior to consent being obtained allowing participants and families consideration time. A further pivot was to involve storying in consent processes. Stories are important for Indigenous peoples as they “spring forth from a holistic epistemology and are the relational glue in socially interdependent knowledge system’^24, p, 108^ Storying involves varying combinations of visual, audio and kinaesthetic methods. For instance, a video of an Aboriginal child having a cardiac MRI, photos of an imaging department, an interactive toy MRI scanner and colouring in sheets designed by local children. Additionally, artwork by an Aboriginal artist assists to tell the research story on consent forms and other research documentation (see Figure 5).

**Figure 5.**
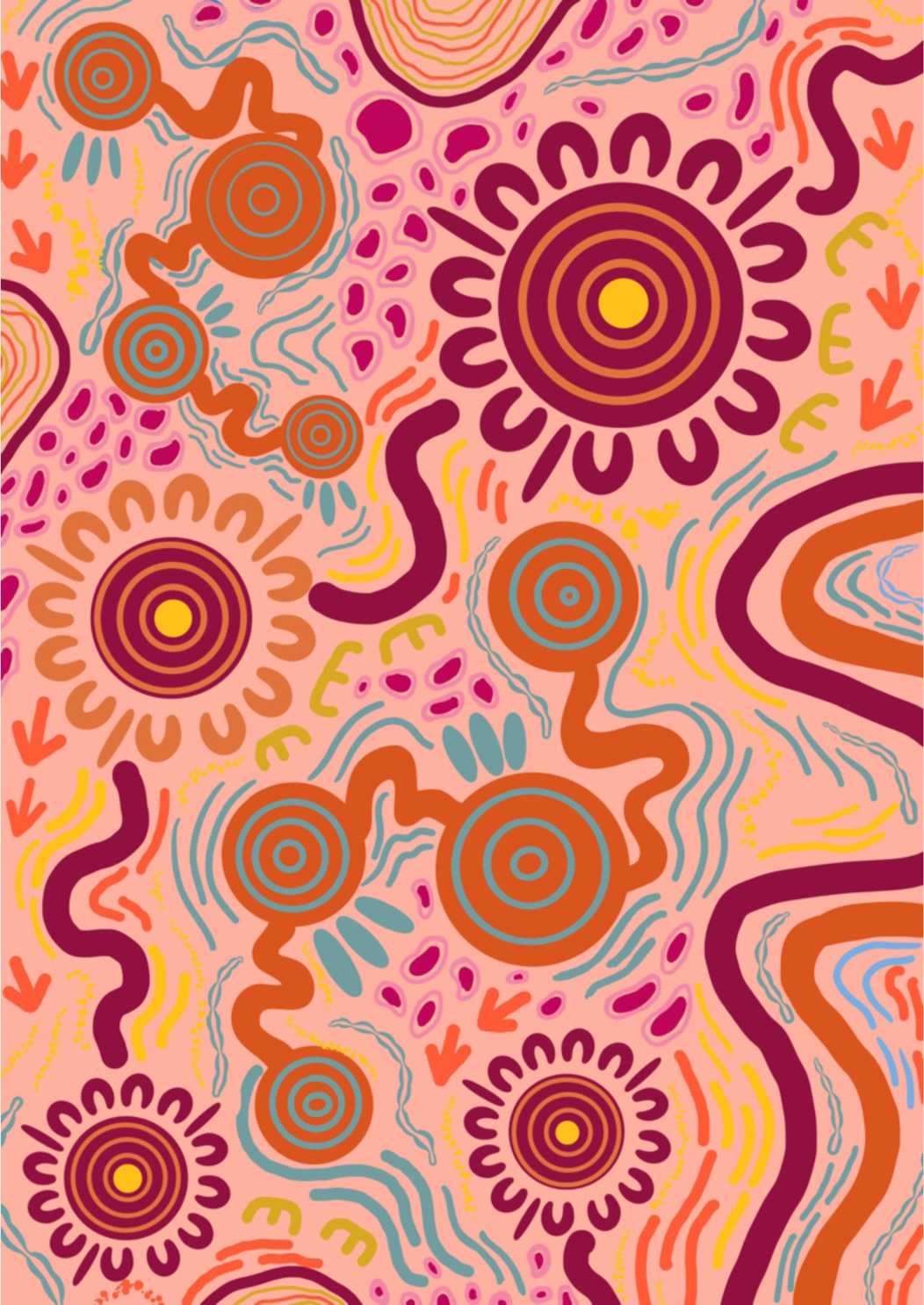
**Journey of Healing, Chloe Jones, Dungala Creations** The main element used in this piece is water. Water is considered a healing agent in many Aboriginal cultures. Water gives life and heals life. The water feature in this piece is the centre part which shows the connection of waterholes through waterways and rivers. The meeting grounds symbolize the merging of our communities to support and heal together. The animal tracks signify the journey of healing by showing a journey along the river. The warm and cool contrast of the colours is also used to show the process of healing. The lined pattern on the edging and the centre represents water or running water and this also represents a journey. Chloe Jones

### Pivot four – multi-site approach

#### Perceived research paradigm clash

A biomedical approach typically involves the project being conducted in a uniform manner at each of the different sites, with a consistent research team structure and hierarchy. An Indigenous Research Paradigm honours the diversity of Country and peoples at local research sites.

#### Responsive pivots

We relied upon engaging local First Nations knowledges at each of the three tertiary hospital settings to responsively shape project conduct, research team structure, recruitment and follow-up processes. Therefore, each site has a unique project structure evolving responsively to local needs. There were, however, consistencies present for project validity, for example, the cardiac MRI protocol and data collection points. Importantly, a collaborative non-hierarchical team approach ensued where local First Nations knowledges were amplified (see Figure 6). This led to the development of site-specific resources, such as certificates of participation and an activity book. The latter aims to reduce boredom during admission, to build familiarity with the hospital and MRI scanner and to create a sense of belonging through activities involving local animals and bush medicines. Furthermore, at the Darwin site, healthy control participants are scanned in small groups on weekends. The families come together to await the scans, and during this time children can play with model scanners, watch the project video and ask questions of the First Nations researchers. This aims to build a sense of community and overcome the discomfort that many First Nations peoples feel in a hospital environment.

**Figure 6.**
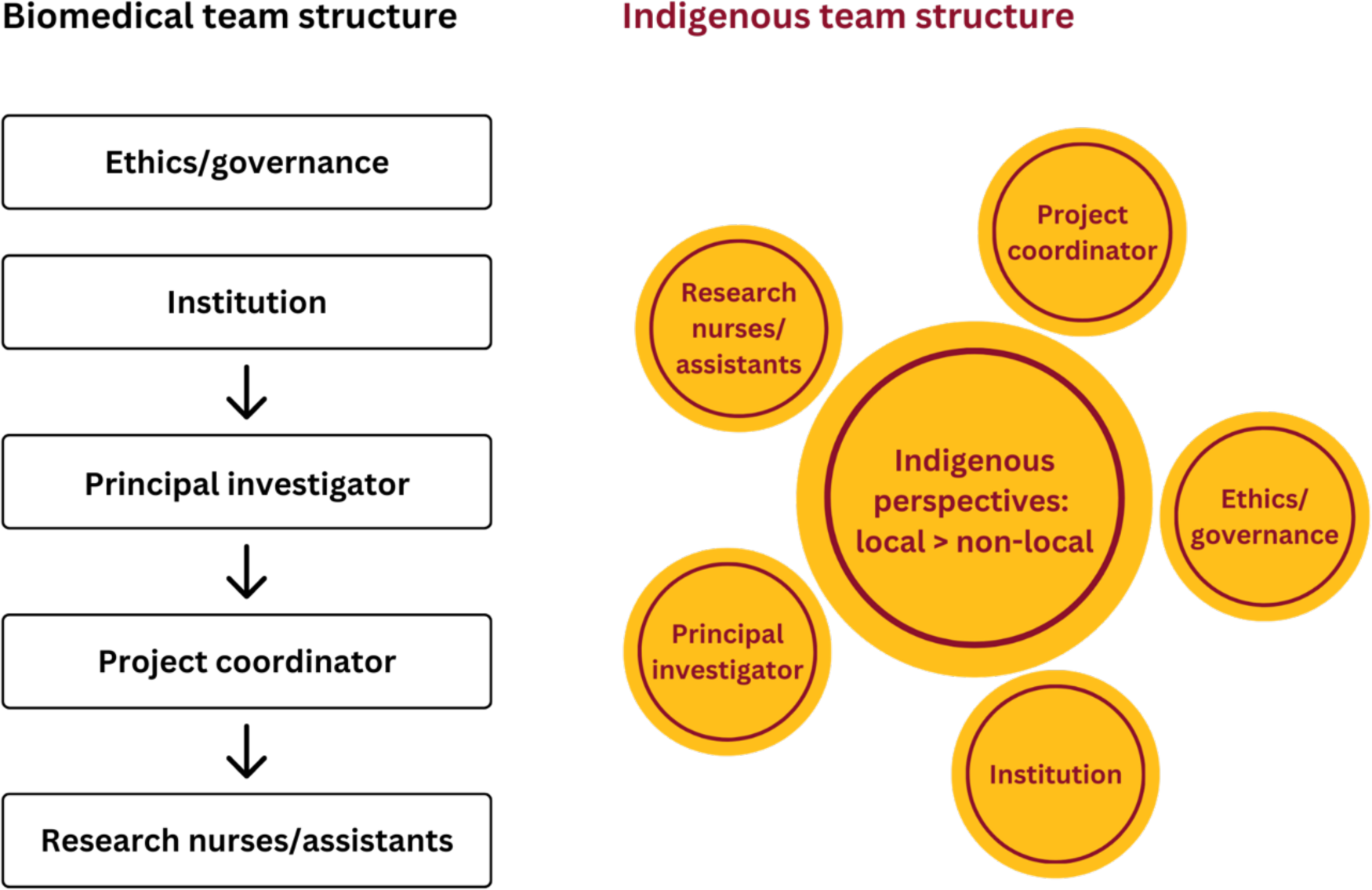
**Research team structure** The biomedical research team structure is typically hierarchical whereas an Indigenous Research team privileges Indigenous perspectives (particularly local), regardless of position or role.

## Discussion

Here we applied Yarning method to reflect on and describe inclusion of an Indigenous Research Paradigm into the establishment of a prospective multi-site tertiary healthcare study on improving diagnosis and treatment of ARF. Our findings demonstrate that principles of an Indigenous Research Paradigm ^1,9^ can be applied in prospective biomedical research in tertiary healthcare institutions. Crucial to the study design was to responsively pivot from a biomedical to Indigenous Research Paradigm. These responsive pivots involved stakeholder engagement, research design, consent processes and multi-site approach.

Critical to the study establishment was multi-layered First Nations leadership. Firstly, having an Aboriginal research lead increased relational accountability. For instance, if the lead created negative research consequences the researcher would likely be held accountable through complex and far-reaching family and kinship connections. The Aboriginal research lead also shared diverse tacit and explicit commonalities with First Nations stakeholders creating immediate commonality for relationality. Particularly shared was a personal investment in First Nations’ futures. Notably, some research participants were surprised to learn the lead researcher was an Aboriginal doctor as they did not realise First Nations doctors existed. Further, First Nations leads included Aboriginal Health Workers, Liaison Officers, cardiac nurses, other First Nations leaders in cardiac health, CCOs and, families affected by ARF. These leads were key to informing responsive pivoting with multiple benefits. For instance, improving the research design and consent processes. While this leadership has led to establishment and implementation of this study, we acknowledge that its ability to assure cultural safety for participants has yet to be determined.

There were several barriers to applying Indigenous Research Paradigm principles in this ARF study. First are medical research training programs lacking inclusion of Indigenous Research Paradigm principles alongside a provision of dominant training in a biomedical paradigm. For example, author one, while Aboriginal, had predominantly trained in a biomedical informed system. This led to indoctrination into a western biomedical model with a subsequent lack of understanding of how to apply Indigenous paradigms in research and clinical settings. This sub-optimal training likely contributes to why biomedical researchers can struggle to work in Indigenous contexts.^28^

A second barrier involves research being conducted in environments where provision of culturally safe clinical care is not guaranteed, particularly with increasing incidences of First Nations peoples experiencing racism during hospital admissions.^29^ In these vulnerable circumstances, coercion and fear can interfere with the ability to provide genuine informed consent. A final barrier involves system failures to support implementation of an Indigenous Research Paradigm. For instance, biomedical research systems and resourcing often focus on time-orientated, linear, and inflexible processes.^30^ These can be unsuited to relational based research methodologies and studies simultaneously attempting to improve healthcare equity.^30^ Problematically, these barriers outlined above will also be contributing to the exclusion of Indigenous peoples in clinical trials and other intervention-based research.^28^

In conclusion, biomedical research undertaken in institutional settings can be altered to become more aligned with an Indigenous Research Paradigm’s principles provided the research is Indigenous led, utilises Indigenous and decolonising methodologies and is informed by multi-layered local Indigenous perspectives. Biomedical researchers must commit to practicing reflexivity and learning about research paradigms outside of the biomedical.

## Contributors

JO led establishment of the prospective “I Heart MRI” research project over a three-year period, participated in the Yarning and contributed to manuscript writing. KA contributed to the conceptual framework, the Yarning and writing of the manuscript. SJG, VW, TW and AJT contributed to design and establishment of “I Heart MRI” and manuscript writing.

The “I Heart MRI” research team includes Ms Toni Walker, Ms Linda Okwaro, Ms Vicki Wade, Dr Bo Remenyi, Ms Erin Ferguson, Ms Alicia Sewcharran, Dr Benjamin Reeves, Ms Melissa van Leeuwen, Dr Angus Baumann, Dr Sarah J Gutman, Dr Jessica O’Brien, and Professor Andrew J Taylor (PI).

## Abbreviations

ARF: acute rheumatic fever
CBPR: community-based participatory research
CCO: community-controlled organisations
HREC: human research ethics committee
MRI: magnetic resonance imaging
RHD: rheumatic heart disease

## Supporting information

I Heart MRI study protocol

## Data Availability

All data produced in the present work are contained in the manuscript

