## Supplementary material for "Taking on colonial institutions: making room for an Indigenous Research Paradigm in prospective biomedical research in tertiary hospitals": I Heart MRI study protocol

**Study protocol for I Heart MRI: Quantifying myocardial inflammation in acute rheumatic fever and rheumatic heart disease**

Jessica O'Brien[a,b,c], Sarah J Gutman[a,b,d], Toni Walker[e], Vicki Wade[e], Karen Adams[c], Andrew J Taylor[a,b,d,f]

a Department of Cardiology, Alfred Hospital, Melbourne, Victoria, Australia
b School of Translational Medicine, Monash University, Melbourne, Victoria, Australia
c Indigenous Health Unit, School of Medicine, Nursing and Health Sciences, Monash University, Clayton, Melbourne, Victoria, Australia
d Baker Heart and Diabetes Institute, Melbourne, Victoria, Australia
e Global and Tropical Health Division, Menzies School of Health Research, Darwin, Northern Territory, Australia
f Department of Cardiology, Royal Melbourne Hospital, Melbourne, Victoria, Australia

**Word count:**

**Funding:** National Health and Medical Research Council (NHMRC) and Heart Foundation First Nations CVD grant

### Introduction

Acute rheumatic fever (ARF) is a preventable cause of heart disease in Australia, disproportionately affecting First Nations children and young people. [1] This pattern of high rates in First Nations peoples with near eradication in the non-Indigenous population is seen in other colonised high-income countries. [2,3] It demonstrates the enduring effects of colonisation, in particular, socioeconomic disadvantage, high density living, inadequate access to acceptable healthcare, all of which are associated with the development of ARF [4,5] and therefore its long-term sequela, rheumatic heart disease (RHD).

ARF diagnosis is based on a combination of clinical manifestations and evidence of preceding streptococcal infection, first outlined by Jones in 1944.[6] Australia has developed its own version of these guidelines[7] after strict application of the Jones criteria led to missed diagnoses due to mild or atypical presentations. [8] Despite diagnostic criteria being clarified as much as practicable, over 40% of diagnoses continue to fall into “possible” or “probable” categories, a number that continues to rise.[9,10] Accurate categorisation is crucial as it guides the duration of secondary antibiotic prophylaxis (SAP). This painful monthly intramuscular injection with benzathine benzylpenicillin G (BPG) is the most efficacious means of preventing ARF recurrence and the subsequent development of RHD.[11] However, it poses significant challenges for patients, families, communities and the health care system, demonstrating the importance of judicious prescribing. Traumatic experiences as well as racism and discrimination at individual and institutional levels are commonly reported by children and young people receieving SAP. [12] Negative experiences with the healthcare system are particularly problematic with long-reaching consequences for First Nations peoples in

Australia where higher rates of coronary artery disease, stroke, diabetes, renal disease, cancers and many other chronic conditions are seen. [13,14]

A major criterion for ARF diagnosis is carditis, currently defined as either clinical (significant murmur, cardiac enlargement, cardiac decompensation and pericardial friction rub or effusion) or subclinical (evidence of valvulitis on echocardiogram). [7] However, not all cardiac involvement is evident on echocardiography during the early stages of ARF, such as during the acute admission. Australian data has consistently shown that more patients with ARF had prolonged PR interval (a minor manifestation) recorded than clinical or subclinical carditis, [9,15] suggesting subtle cardiac involvement despite normal echocardiogram findings. The delayed development of macroscopic structural and functional changes is especially disruptive if patients have already been discharged with SAP plans in place, particularly if they are geographically distant from the echo services and medical specialists required for diagnostic revision.

Aschoff bodies are the pathognomonic myocardial biopsy finding in ARF. [16] They are interstitial fibroinflammatory lesions made up of various inflammatory substances including lymphocytes, macrophages, B cells and Giant cells. [17] While associated with collagen necrosis and interstitial oedema, they do not appear to involve myocyte necrosis,[18] and as such, multiple studies have demonstrated normal high sensitivity troponin concentrations even in ARF patients with confirmed carditis. [18-23] Elevated troponins have been documented, but typically when severe valvular dysfunction is present and complicated by decompensated heart failure, [19] an uncommon presentation of ARF. Thus, high sensitivity troponin offers limited value as a diagnostic tool. However, this interstitial inflammatory infiltrate represents a potential target for diagnosis, provided it can be identified with a non-invasive approach suitable for children and young people.

Non-contrast T1 mapping, a sequence available on cardiac magnetic resonance imaging (CMR) has proven highly accurate in the detection of inflammatory infiltrate in the setting of acute myocarditis. [24] Unfortunately, widespread clinical uptake has been impeded by the variety of sequences and analysis techniques used by different institutions, making comparison across institutions difficult, and resulting in a paucity of robust research in this area. The most recent position statement on the clinical application of CMR mapping techniques suggested that T1 indices should be presented as Z-scores (the number of standard deviations by which the result differs from the local normal reference mean) to overcome differences between local reference T1 values. [25] This approach has been used successfully, with conversion of raw T1 values to Z-scores markedly improving the agreement between T1 mapping techniques. [26]

CMR is only available in metropolitan areas or large regional centres, far from the remote communities with the highest rates of ARF,[1] however, in Australia when children or young people are suspected to have ARF they are transferred to the nearest tertiary hospital for further investigation and assessment by a specialist team.

Our group conducted a pilot study at Alice Springs hospital from 2017 to 2019, assessing non-contrast T1 time on CMR undertaken during the admission.[27] It showed that children and young people with probable or definite ARF had higher non-contrast T1 times compared with inflammatory and healthy controls, even when electrocardiogram and echocardiography was normal. This suggests that T1 time can be utilised to identify rheumatic carditis and therefore clarify the presence of ARF in those patients where diagnosis is uncertain.

Primary objectives

The primary objectives of this study are:

1. To develop a cross-institutional score for the diagnosis of ARF based on non-contrast T1 time.
2. To determine the ability of CMR imaging to predict the future development of RHD in people presenting with ARF.

Importantly, these research objectives meet the needs identified by the communities most impacted by ARF and RHD, namely, clarification of the need for SAP and identification of those children and young people at highest risk of developing RHD.

### Methods

Study design and methodology

Quantifying myocardial inflammation in ARF and RHD or I Heart MRI is a multi-site prospective observational study. Children and young adults with definite, probable or possible ARF (either initial or recurrent episodes) and those with non-ARF inflammatory conditions (such as septic arthritis or pneumonia) will be prospectively recruited. A local non-contrast T1 reference standard will be developed from the T1 times of 15 healthy controls at each participating site. Both inflammatory and healthy control participants will be matched by age, sex and ethnicity.

All participants will undergo the same non-contrast truncated CMR scan. For ARF and inflammatory control participants troponin will be undertaken on previously collected blood if not already performed; venesection will not be repeated (the utility of this additional information is not anticipated to outweigh the burden on participants of a needle-associated intervention). There is no change to routine investigation or management; the need for, and duration of, SAP will remain at the discretion of the treating team.

Participants with ARF will be followed up at two to three years by telephone interview to determine if any (known) ARF recurrences have occurred or if a diagnosis of RHD has been given. It will also allow the opportunity to directly feedback aggregate results from baseline scans and give a general update on project progress. Where participants cannot be contacted, information regarding recurrences, SAP and progression to RHD (as determined by transthoracic echocardiogram) will be obtained from the Northern Territory and Queensland RHD Program registers. Consent to access their register data will be obtained as part of the initial consent process. Participants will be invited back for repeat CMR to determine if T1 time remains elevated even in the absence of known ARF recurrences and progression to RHD.

This flexible follow-up timeframe reflects that clinical echocardiograms will be used to determine progression to RHD rather than necessitating return to a tertiary hospital for a research echocardiogram. If a clinically indicated echocardiogram has not been undertaken in the 12 months prior to follow-up, it will be arranged by the research team but undertaken as close to participants' homes as possible (for example, during outreach visits where tertiary healthcare teams go to communities to provide care). This timeframe is also informed by risk of progression to RHD which is highest in the first year after ARF diagnosis then decreases thereafter.[28]

Given that this patient population is young, predominantly First Nations peoples and a group who rely on frequent engagement with health services to prevent RHD, robust project design has been balanced against avoiding interference with RHD education and uptake of SAP to mitigate negative effects of participation in this research.

Study sites

There are three participating hospitals across two states/territories in Australia; Darwin (Larrakia Country) and Alice Springs (Arrernte Country), both in the Northern Territory (NT), and Cairns hospital (Gimuy Walubara Yidinji & Yirrangangi Country) in Queensland. These sites were selected as they have MRI scanners capable of undertaking CMR as well as high numbers of people being admitted with ARF.

Study participants

Eligible patients, aged between eight and 35 years of age, are identified by their treating team during inpatient admission with definite, probable or possible ARF. Inflammatory control participants are similarly recruited from the wards. Inclusion and exclusion criteria are presented in Table 1.

Most participants are expected to be First Nations peoples, so significant diversity in language, culture and ways of living is anticipated. All aspects of project design and conduct have been developed with this in mind, informed by local First Nations stakeholders at each site.

Ethics and informed consent

During 2020-2021 this project was approved by the Far North Queensland Human Research Ethics Committee (HREC) and the Human Research Ethics Committee of the Northern Territory Department of Health and Menzies School of Health Research. The project is conducted in accordance with the Australian Institute of Aboriginal and Torres Strait Islander Studies (AIATSIS) Code of Ethics for Aboriginal and Torres Strait Islander Research,[29] as well as the National Health and Medical Research Council (NHMRC) ethical guidelines for research with Aboriginal and Torres Strait Islander peoples.[30] It has been prospectively registered with the Australian New Zealand Clinical Trials Registry (ACTRN12621001463864).

Written informed consent is obtained from participants or their guardian if aged under 18 years. Per the Far North Queensland HREC, a written assent form is also used at the Cairns site. It states that the guardian believes the participant is willing to be involved in the project.

Each participant and family are offered the involvement of an Aboriginal and Torres Strait Islander Liaison Officer or Health Worker and where applicable, an interpreter. If an interpreter is required but unavailable, no attempts at recruitment are made. In keeping with an Indigenous paradigm, involving other family and community members in the consent discussion is encouraged. Broader and long-term effects of study participation, including potential effects on family and community are discussed. Wherever possible, a First Nations researcher obtains consent.

Research team

The research team includes four First Nations researchers (JO, VW, TW, KA). Non-Indigenous team members include CMR experts (AJT, SJG) and clinician researchers with extensive experience in Indigenous health, in particular, with ARF and RHD.

First Nations advisory group

In addition to this project being First Nations-led, a First Nations advisory group oversees study conduct. The group consists of seven members, all First Nations peoples with experience in First Nations health and research. The group receives updates on the project and provide feedback about progress and any issues raised. Advisory group opinions will also inform the dissemination of results to participants and the broader First Nations community, though local First Nations perspectives will provide the foundation of these site-specific resources.

CMR protocol

CMRs at Darwin hospital will be undertaken on a 1.5T Philips Ingenia, at Alice Springs hospital on a 3-T Siemens MAGNETOM Skyra and at Cairns hospital on a 3-T Siemens MAGNETOM Vida.

The protocol is truncated and non-contrast, designed to make the process more suitable for children and young people. After acquisition of scout images, mid short axis T1 and T2 mapping is undertaken prior to cine imaging in standard 4-, 3-, and 2-chamber long-axis views using an ECG-gated balanced steady-state free precision (SSFP) sequence in expiration. Short tau inversion recovery (STIR) imaging follows if time permits. If a participant is unable to perform breath holds, free breathing sequences are performed. The unconventional order of sequences is to prioritise those that are the focus of the project in case a participant wishes to end the scan prematurely.

Calculation of LV volume and function is undertaken via biplane analysis using CMR42 (Circle Cardiovascular Imaging Inc., Calgary Canada). T1 mapping is performed with images processed via a curve fitting technique to generate T1 maps for calculation of the non-contrast myocardial T1 time. This will be performed by two cardiologists, trained in cardiac MRI, blinded to clinical data.

Endpoint measures

The primary endpoint is T1 time Z-score of definite ARF participants compared to non-ARF inflammatory control participants. Secondary endpoints include:

- progression to RHD at two to three years (as determined by transthoracic echocardiogram) in the possible and probable ARF group, adjusting for doses of BPG received
- T2 time Z-score
- STIR ratio
- difference in T1 time at follow-up compared to baseline (in the subgroup able to return for repeat CMR).
- High sensitivity troponin

Data collection and management

Baseline and outcome data will be stored and collated into a standalone REDCap database. Local research team members only have access to the data of participants at their site. The First Nations project lead (JO) is the only research team member to have access to the identifiable data of all project participants.

Data collected includes: demographics, date of symptom onset, clinical presentation including ARF manifestations, laboratory results including troponin, diagnosis assigned by the medical team, echocardiography results and follow-up planned on discharge. Diagnosis of covid or administration of covid vaccine in the six weeks prior to enrolment is documented as both have been associated with elevation of T1 time when myocarditis is present.[31,32]

### Assignment of diagnosis

The research team will compare each participant's final diagnosis upon hospital discharge with their clinical, laboratory and echocardiographic information. Where a diagnosis appears inconsistent with the 2020 Australian guidelines for the prevention, diagnosis and management of acute rheumatic fever and rheumatic heart disease,[7] the case will be adjudicated by two clinicians highly experienced in ARF.

### Sample size and statistical analysis

A reference standard, from which a Z score for healthy myocardium can be developed, will be determined at each site based on the T1 times of 15 healthy controls. Patients with definite ARF based on the 2020 Australian ARF/RHD guidelines will also be enrolled at each centre, and the measured non-contrast T1 times of ARF patients will be compared against the local reference Z-score and the Z-scores of the non-ARF inflammatory controls.

Based on the data from the Alice Springs pilot study, in which control participants had T1 times of 821 ± 57ms, a patient with a non-contrast myocardial T1 Z-score of 2 (2 standard deviations above the local reference mean) would have a measured non-contrast T1 of 935ms (i.e. 821 + 2 times 57ms). Utilizing a non-contrast myocardial T1 Z-Score >2 as the threshold for the diagnosis of carditis from ARF, a total of 40 patients with ARF pooled across all institutions will result in a statistical power of greater than 95% at a significance level of P<0.05. In addition, we will perform receiver operating characteristic curve analysis to determine the optimal cut-off for the myocardial z-score to diagnose ARF.

To test the hypothesis that patients with possible or probable ARF are more likely to progress to RHD if their non-contrast T1 time was elevated at the time of diagnosis, echocardiography will be used to identify progression to RHD at follow up. Patients will be dichotomised into those with normal and those with elevated non-contrast myocardial T1 time. Utilizing the log rank test for equality of survival curves, a total of 30 patients will be required in each group for a statistical power of 86% to demonstrate an incidence of at least 15% for the development of RHD in those with an elevated non-contrast myocardial T1 time, assuming those with a normal myocardial T1 time do not develop RHD. Given we will not know which patients have an elevated non-contrast T1 time at enrolment, we expect we will need to recruit 80 patients with possible or probable ARF in order to have 30 with an elevated T1 time.

**Discussion**

This project aims to determine whether CMR can identify subtle subclinical rheumatic carditis in people with a range of ARF presentations, improving diagnostic accuracy and therefore clarifying whether an individual will benefit from SAP.

Based upon the Australian ARF and RHD guidelines, SAP duration is the same for probable and definite ARF (five years or until age 21, whichever is longest) provided there is no evidence of RHD on echocardiography.[7] However, data from the Northern Territory register shows a significantly higher number of BPG doses received in those with a definite diagnosis of ARF compared with probable,[9] suggesting that certainty of diagnosis has an effect on patients and families' engagement with the SAP program. Efficacy of SAP is strongly linked to the percentage of doses received, with greatest benefit achieved at ≥ 80% BPG doses administered.[33] If non-contrast T1 time Z score can demonstrate the extent to which an individual will receive benefit from SAP, it is likely to improve engagement with SAP and therefore reduce the risk of ARF recurrence and progression to RHD.

Conversely, if CMR shows that a T1 Z score close to 0 is associated with an extremely low risk of progression to RHD, it may identify a group of children and young people who are receiving little, if any, benefit from SAP and therefore could be monitored closely without it. The low rates of progression to RHD seen in the possible ARF cohort (2% in a recent analysis of Northern Territory register data) [9] likely reflect that the group includes those who never had ARF, and perhaps also those who have a form of ARF that confers very low risk of RHD. T1 Z score will need to be interpreted within the context of other available information, and will not serve as a single diagnostic test for ARF. However, with informed decision-making from children and young people, their families and trusted community health services, T1 Z score could allow for individualised approaches to follow-up and SAP plans. This approach aligns well with the current development of programs involving community-based handheld echocardiography to monitor children and young people with prior ARF. Health workers obtain images in remote and regional clinics and transfer them securely to cardiologists for reporting. This decentralisation of ARF follow-up will allow for the provision of culturally responsive care with enhanced frequency of echocardiographic monitoring.

I Heart MRI also aims to expand our understanding of the pathophysiology of ARF, in particular, the presence and relevance of subclinical rheumatic carditis. The evidence base examining the role of CMR in the assessment and management of RHD is increasing. However publications regarding CMR in ARF are limited to case reports[34,35] and a study specifically assessing late gadolinium enhancement to identify rheumatic carditis.[36] Investigations requiring needles have consistently been identified by people with ARF and their families as problematic, highlighting that a non-contrast alternative would be greatly valued by this cohort.

The limitations of this study are characteristic of ARF research and result predominantly from the heterogeneity of ARF with regards to clinical phenotype (arthritis, carditis, chorea), diagnostic category (definite, probable, or possible) and whether a presentation is initial or recurrent.[37] Furthermore, scanning participants early or at the same point in the disease process will be challenging due to varying time from symptom onset to hospitalization, time required to obtain culturally safe informed consent and accessing scanner time on busy clinical magnets. That CMR is currently only undertaken in urban and regional institutional settings may impede the translation of study results to clinical practice for adults, who are not routinely admitted to hospital for ARF workup like their paediatric counterparts. However, mobile MRI networks designed to service regional and remote areas are under development, including for other acute conditions such as stroke.

In conclusion, this study will examine whether a CMR-based diagnostic score for ARF can be developed to clarify ARF diagnosis and predict who is at highest risk of progressing to RHD. In doing so, it addresses an important research priority of numerous First Nations communities in Australia.

**Table 1. Inclusion and exclusion criteria**

| Group | Inclusion criteria (all must be present) | Exclusion (if any of the following present) |
|---|---|---|
| ARF | Aged 8 to 35 years<br>Definite, probable or possible ARF | Claustrophobia<br>Metal implant/object that precludes CMR<br>Pregnant or breastfeeding |
| Inflammatory controls | Aged 8 to 35 years<br>Acute inflammatory/infective condition | Any major criteria for ARF<br>History of ARF or RHD<br>Chest pain<br>Signs or symptoms of heart failure<br>More than mild valvular disease on echocardiogram<br>Inflammatory condition commonly caused by Streptococcus where no alternate organism has been identified<br>Known streptococcal infection within 6 months of enrolment<br>Claustrophobia<br>Metal implant/object that precludes CMR<br>Pregnant or breastfeeding |
| Healthy controls | Aged 8 to 35 years | Any infective or inflammatory condition<br>Any major or minor criteria for ARF<br>History of ARF or RHD<br>Chest pain<br>Signs or symptoms of heart failure<br>More than mild valvular disease on echocardiogram<br>Known streptococcal infection within 6 months of enrolment<br>Claustrophobia<br>Metal implant/object that precludes CMR<br>Pregnant or breastfeeding |
